# Incidence Rates of Medically Attended COVID-19 in Infants Less than 6 Months of Age

**DOI:** 10.1101/2022.09.28.22280464

**Authors:** Isabel Griffin, Stephanie A. Irving, Carmen Sofia Arriola, Angela P. Campbell, De-Kun Li, Fatimah S. Dawood, Caroline Doughty-Skierski, Jeannette R. Ferber, Nickolas Ferguson, Louise Hadden, Jillian T. Henderson, Mary Juergens, Venkatesh Kancharla, Allison L. Naleway, Gabriella Newes-Adeyi, Erin Nicholson, Roxana Odouli, Lawrence Reichle, Mo Sanyang, Kate Woodworth, Flor M. Munoz

## Abstract

**Objective:** Studies suggest infants may be at increased risk of severe COVID-19 relative to older children, but few data exist regarding the incidence of COVID-19 episodes and associated risk factors. We estimate incidence rates and describe characteristics associated with medically attended COVID-19 episodes among infants younger than 6 months of age.

**Methods:** We analyzed electronic medical record data from a cohort of infants born March 1, 2020‒ February 28, 2021. Data from three health care delivery systems included demographic characteristics, maternal and infant outpatient visit and hospitalization diagnoses, and SARS-CoV-2 test results. Medically attended COVID-19 episodes were defined by positive SARS-CoV-2 clinical tests and/or COVID-19 diagnosis codes during medical care visits. Unadjusted and site-adjusted incidence rates by infant month of age, low and high SARS-CoV-2 circulation periods and maternal COVID-19 diagnosis were calculated.

**Results:** Among 18,192 infants aged <6 months whose mothers received prenatal care within the three systems, 173 (1.0%) had medically attended COVID-19 episodes. Incidence rates were highest among infants aged under 1 month (2.0 per 1,000 person-weeks) and 1 month (2.0 per 1,000 person-weeks) compared with older infants. Incidence rates were also higher for infants born to women with postpartum COVID-19 compared with women without known COVID-19 and women diagnosed with COVID-19 during pregnancy.

**Conclusion:** Most medically attended COVID-19 episodes in infants aged <6 months were outpatient care encounters. Infants of women with postpartum COVID-19 had a higher risk of medically attended COVID-19 than infants born to mothers who were diagnosed during pregnancy or never diagnosed underscoring the importance of COVID-19 prevention measures for their household members and caregivers to prevent infections in infants.

**Article Summary:** This report describes incidence rates and characteristics of medically attended outpatient and inpatient COVID-19 episodes among infants aged <6 months during the COVID-19 pandemic.

**What’s Known on This Subject:** Surveillance data and case series suggest that infants aged <1 year may be more likely to be hospitalized with COVID-19 than older children, but few data are available about the risk of medically attended COVID-19 episodes among infants.

**What this Study Adds:** Among infants aged <6 months, few were hospitalized with COVID-19. Incidence rates of medically attended COVID-19 episodes were highest among infants aged ≤ 1 month and among infants of women with COVID-19 during the 6 month post-partum period.

---

Early in the coronavirus disease 2019 (COVID-19) pandemic, surveillance data suggested that COVID-19 was infrequent and mild in children but that hospitalization rates among infants aged <1 year might be higher than among older children.^1^ More recent studies suggest that infants aged less than one year of age primarily experience mild to moderate COVID-19, but represent a disproportionate amount of pediatric COVID-19 associated hospitalizations.^2,3,4,5,6^ However, most published studies to date describing severe acute respiratory syndrome-related coronavirus (SARS-CoV-2) infection among infants are based on convenience samples with small numbers of infants that preclude examining outcomes by age group and lack population denominators that allow an assessment of true community risk. Estimates of incidence rates of medically attended COVID-19 episodes among infants, including outpatient care encounters, could help guide risk communication for expectant and new parents and might inform future public health recommendations for this group.

The Epidemiology of SARS-CoV-2 in Pregnancy and Infancy (ESPI) Network: Electronic Cohort (eESPI) collected electronic medical record data for a retrospective cohort of pregnant persons and their infants who were born during the COVID-19 pandemic. Data about infants were collected up until the date they turned six months old. We analyzed data from the eESPI pregnancy and infant cohort to describe infants <6 months of age with and without medically attended COVID-19 episodes including both hospitalizations and outpatient care encounters (including urgent care and emergency department encounters). Additionally, we estimated incidence rates of medically attended COVID-19 episodes overall and by level of SARS-CoV-2 circulation, site, age group, and maternal COVID-19 diagnosis during pregnancy and the six month postpartum period.

## METHODS

### Study design and population

The eESPI study was conducted at three healthcare delivery systems in the United States: Kaiser Permanente Northwest (Portland, OR), Kaiser Permanente Northern California (Oakland, CA), and Baylor College of Medicine (Houston, TX). Persons were eligible for inclusion in the maternal cohort if they 1) had an estimated delivery date (EDD) from March 1, 2020 through February 28, 2021 excluding women with pregnancy losses that occurred prior to March 1, 2020 and 2) had at least one prenatal care outpatient or telemedicine visit within the respective health system from December 1, 2019 through February 28, 2021. The infant cohort included all live born infants of individuals included in the maternal cohort. This analysis included all infants born from March 1, 2020 through February 28, 2021.

### Data collection

Data were extracted from electronic medical records using standardized variable definitions across study sites (see Supplemental Methods Data Dictionary – Appendix A and Appendix B). Most variables were defined by one or more International Classifications of Diseases 10 (ICD-10) codes. Data collection included maternal and infant demographic characteristics, maternal prenatal visit history, maternal and infant medical conditions, diagnosis codes for all telemedicine, emergency department (ED), and ambulatory care encounters and hospitalizations, and results from all SARS-CoV-2 RT-PCR testing. For this study, data about infants were collected from birth until they turned six months of age or their age as of February 28, 2021, whichever occurred first.

### Analysis definitions

All variables derived from ICD-10 codes can be found in Appendix A and Appendix B. Medically attended COVID-19 episodes in infants, including telemedicine, ED, and ambulatory care encounters, were identified based on at least one of the following: 1) a positive RT-PCR test for SARS-CoV-2 within three days of a medical encounter, or 2) a COVID-19 diagnostic code (ICD-10: U07.1) (Appendix A). Hospitalizations for any reason were counted if they met the test and diagnostic criteria described above.

Maternal race/ethnicity was extracted from electronic medical records and classified as Hispanic/Latina, non-Hispanic White, non-Hispanic Black, and non-Hispanic Other. The non-Hispanic Other race/ethnicity category included American Indian or Alaska Native, Asian, Native Hawaiian or Pacific Islander, and women who had more than one race listed in their medical record. A list of conditions determined to be underlying medical conditions for mothers and infants can be found in the Supplemental Methods Data Dictionary (Appendix B). Timing of underlying medical conditions for infants was based on timing relative to any COVID-19 diagnosis or positive test. Timing of COVID-19 for mothers was determined based on either a documented positive RT-PCR or antigen test for SARS-CoV-2 or COVID-19 diagnostic code during pregnancy or during the postpartum period (two mothers had COVID-19 documented during both pregnancy and the postpartum period). The postpartum period was defined as the six months immediately following delivery.

Gestational age at delivery (preterm [<37 weeks] or term/post-term [≥37 weeks]) was determined using reported gestational age at delivery or derived from delivery date and estimated date of conception (Appendix A). High and low SARS-CoV-2 circulation weeks were defined for each site as weeks with case counts above the median case count (high) or below the median case count (low) for the study period, using Health and Human Services (HHS) representative county-level surveillance data for Texas (Harris County), Oregon (Multnomah County), and California (Santa Clara County) in each health system’s service area.^7^

### Analysis

Frequencies of maternal and infant characteristics were described for infants with and without medically attended COVID-19 episodes. Available characteristics of interest included maternal race/ethnicity, maternal Medicaid status, maternal COVID-19 diagnosis (during pregnancy, during the six month postpartum period, or both during pregnancy and the postpartum period), gestational age at birth, infant sex, presence of any underlying medical conditions for mother and infant, and site. Unadjusted chi-square and fisher’s exact test p-values of ≤ 0.05 were considered statistically significant.

For incidence rate calculations, an aggregate crude incidence rate for the full cohort was calculated with the total number of infants with medically attended COVID-19 episodes as the numerator and the total number of person-weeks at risk that each infant (those with and without medically attended COVID-19 episodes) was in the cohort as the denominator. Site-adjusted negative binomial models were then used to estimate incidence per 1,000 person-weeks and 95% confidence intervals by infant sex, maternal race/ethnicity, maternal Medicaid status, presence of any underlying medical conditions for mother and infant, age in months (<1 month, 1 month, 2 months, 3 months, 4 months, and 5 months of age), maternal SARS-CoV-2 infection status (no infection, infection during pregnancy or infection during the six-month postpartum period), and low or high SARS-CoV-2 circulation in the community.

All analyses were conducted in SAS Version 9.4.

### Ethical Review

This protocol was reviewed by the Institutional Review Boards of participating sites. This activity was reviewed by CDC and was conducted consistent with applicable federal law and CDC policy.

## RESULTS

Among 18,192 infants aged less than six months of age in the cohort, 173 (1.0%) had medically attended COVID-19 episodes, including 114 (65.9%) identified by positive SARS-CoV-2 laboratory testing and 59 (34.1%) by COVID-19 diagnosis code only without a documented laboratory-test in the medical record. Medically attended COVID-19 episodes were more frequent among infants from site B (1.4% of 7,434 infants) compared with those from sites A and C (site A 0.8% of 4,794 infants and site C 0.5% of 5,964 infants) (Table 1). There were no differences between infants with versus without medically attended COVID-19 episodes by infant sex or gestational age at birth. However, compared to infants without medically attended COVID-19, a larger proportion of infants with medically attended COVID-19 episodes were infants of Hispanic mothers (46.8% vs. 27.9%, <0.0001), Medicaid beneficiaries (29.5% vs. 22.4%, p=0.05), and mothers with postpartum COVID-19 (57.2% vs. 1.5%, p≤0.0001). Of the 173 infants with medically attended COVID-19 episodes, 27 (15.6%) were diagnosed at <1 month of age, 29 (16.7%) at 1 month of age, 28 (16.2%) at 2 months, 16 (9.2%) at 3 months, 31 (17.9%) at 4 months, and 42 (24.3%) at 5 months of age. A larger proportion of infants with medically attended COVID-19 episodes also had any underlying medical condition (12.1% vs. 7.3%, p=0.02). Six of 173 infants with medically attended COVID-19 (3.5%) had an underlying heart-related condition, 11 (6.4%) had an underlying lung-related condition, and 1 (0.6%) had a liver-related condition (Appendix A); all conditions were documented prior to or on the same days as the infant’s COVID-19 diagnosis or positive SARS-CoV-2 test result.

**Table 1.** Maternal and Infant Characteristics by whether Infants had Medically Attended Coronavirus Disease (COVID-19) Episodes before 6 Months of Age, Epidemiology of SARS-CoV-2 in Pregnancy and Infancy (ESPI) Network: Electronic Cohort (eESPI) (N= 18,192 infants).

|  | Total<br>(N= 18,192) | Infants with<br>Medically<br>Attended<br>COVID-19<br>Episodes<br>*n*<br>(n=173) | Infants without<br>Medically<br>Attended<br>COVID-19<br>Episodes<br>*n*<br>(n=18,019) | *P*<sup>a</sup> |
| --- | --- | --- | --- | --- |
| Characteristics |  | *n* (row %) | *n* (row %) |  |
| *Study Site* |  |  |  | <0.0001 |
| A | 4794 | 38 (0.8) | 4756 (99.2) |  |
| B | 7434 | 106 (1.4) | 7328 (98.6) |  |
| C | 5964 | 29 (0.5) | 5935 (99.5) |  |
| *Infant Sex* |  |  |  | 0.66 |
| Male | 9345 | 93 (1.0) | 9252 (99.0) |  |
| Female | 8842 | 80 (0.9) | 8762 (99.1) |  |
| Unknown | 5 | 0 (0.0) | 5 (100) |  |
| *Maternal Race/Ethnicity*<sup>b</sup> |  |  |  | <0.0001 |
| Non-Hispanic White | 7583 | 32 (0.4) | 7551 (99.6) |  |
| Non-Hispanic Black | 1871 | 25 (1.3) | 1846 (98.7) |  |
| Hispanic | 5112 | 81 (1.6) | 5031 (98.4) |  |
| Other | 3318 | 30 (0.9) | 3288 (99.1) |  |
| Unknown | 308 | 5 (1.6) | 303 (98.4) |  |
| *Maternal Medicaid* |  |  |  | 0.05 |
| Yes | 4085 | 51 (1.2) | 4034 (98.8) |  |
| No | 13947 | 120 (0.9) | 13827 (99.1) |  |
| Unknown | 160 | 2 (1.2) | 158 (98.8) |  |
| *Maternal COVID-19* |  |  |  | <0.0001 |
| *Diagnosis<sup>c</sup>* |  |  |  |  |
| No Known COVID-19 | 17122 | 66 (0.4) | 17056 (99.6) |  |
| During Pregnancy only | 623 | 6 (1.0) | 617 (99.0) |  |
| During Post-partum | 367 | 99 (27.0) | 268 (73.0) |  |
| Period |  |  |  |  |
| During Pregnancy and | 80 | 2 (2.5) | 78 (97.5) |  |
| Post-partum Period |  |  |  |  |
| *Gestational Age at Birth<sup>d</sup>* |  |  |  | 0.31 |
| Preterm (< 37 weeks) | 1819 | 18 (1.0) | 1801 (99.0) |  |
| Term or Post-Term (≥37 | 16373 | 155 (1.0) | 16218 (99.0) |  |
| weeks) |  |  |  |  |
| *Any Underlying Medical Conditions* |  |  |  |  |
| Infant<sup>e</sup> |  |  |  |  |
| Yes | 1327 | 21 (1.6) | 1306 (98.4) | 0.02 |
| No | 16865 | 152 (0.9) | 16713 (99.1) |  |
| Maternal<sup>e</sup> |  |  |  |  |
| Yes | 10320 | 91 (0.9) | 10229 (99.1) | 0.001 |
| No | 7872 | 82 (1.0) | 7790 (99.0) |  |

<sup>a</sup> Unadjusted chi-square and fisher's exact test p-values of $\leq 0.05$ were considered statistically significant.
<sup>b</sup> Race/Ethnicity was extracted from medical records.
<sup>c</sup> Maternal COVID-19 diagnosis was based on a recorded COVID-19 diagnosis (ICD-10: U07.1) or a recorded visit with confirmed SARS-CoV-2 infection by any method (RT-PCR, antigen, etc.) based on hospital records at any time during the pregnancy or during the six months after delivery.
<sup>d</sup> Gestational age at delivery (preterm [ $<37$ weeks], term [ $37-42$ weeks], or post-term [ $>42$ weeks]) was determined using reported gestational age at delivery or derived based on delivery date and estimated date of conception.
<sup>e</sup> Definition of any underlying medical conditions for mothers and infants can be found in Appendix.

Among the 173 infants with medically attended COVID-19 episodes, 148 (85.6%) were first diagnosed in association with telemedicine visits, 20 (11.6%) during ambulatory/outpatient visits, one (0.6%) during an emergency room visit, and four (2.3%) during hospitalization (data not shown). All four hospitalizations occurred among infants aged 0-1 month. Hospital lengths of stay ranged from 2-5 days, and none included intensive care unit admission, mechanical ventilation, or extracorporeal membrane oxygenation.

### Incidence of Medically Attended COVID-19 Episodes

The overall cumulative incidence of medically attended COVID-19 episodes was 0.95%. The aggregate crude incidence rate among the full cohort was 0.6 per 1,000 person-weeks. Site-adjusted incidence rates of medically attended COVID-19 episodes varied by local SARS-CoV-2 COVID-19 circulation (high versus low), infant age, maternal race/ethnicity, maternal Medicaid status, infant underlying conditions, and maternal COVID-19 diagnosis (Table 2). The overall site-adjusted incidence rates of medically attended COVID-19 episodes were 0.6 per 1,000 person-weeks during high SARS-CoV-2 circulation periods and 0.4 per 1,000 person-weeks during low circulation periods. By infant age, the site-adjusted incidence rates of medically attended COVID-19 episode were highest among infants under 1 month and 1 month of age (2.0 and 2.0 per 1,000 person-weeks, respectively) compared with older infants.

**Table 2.**
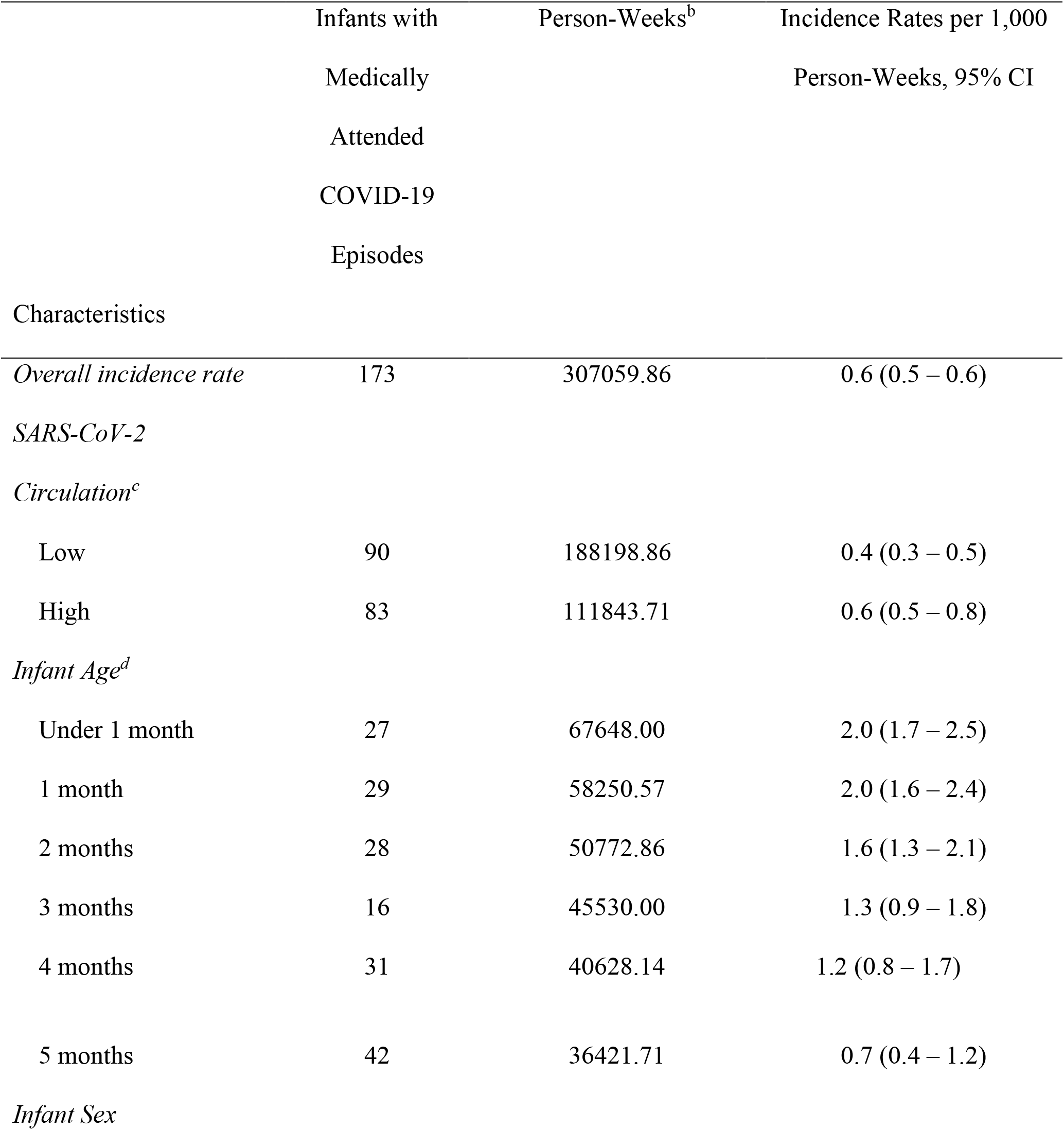
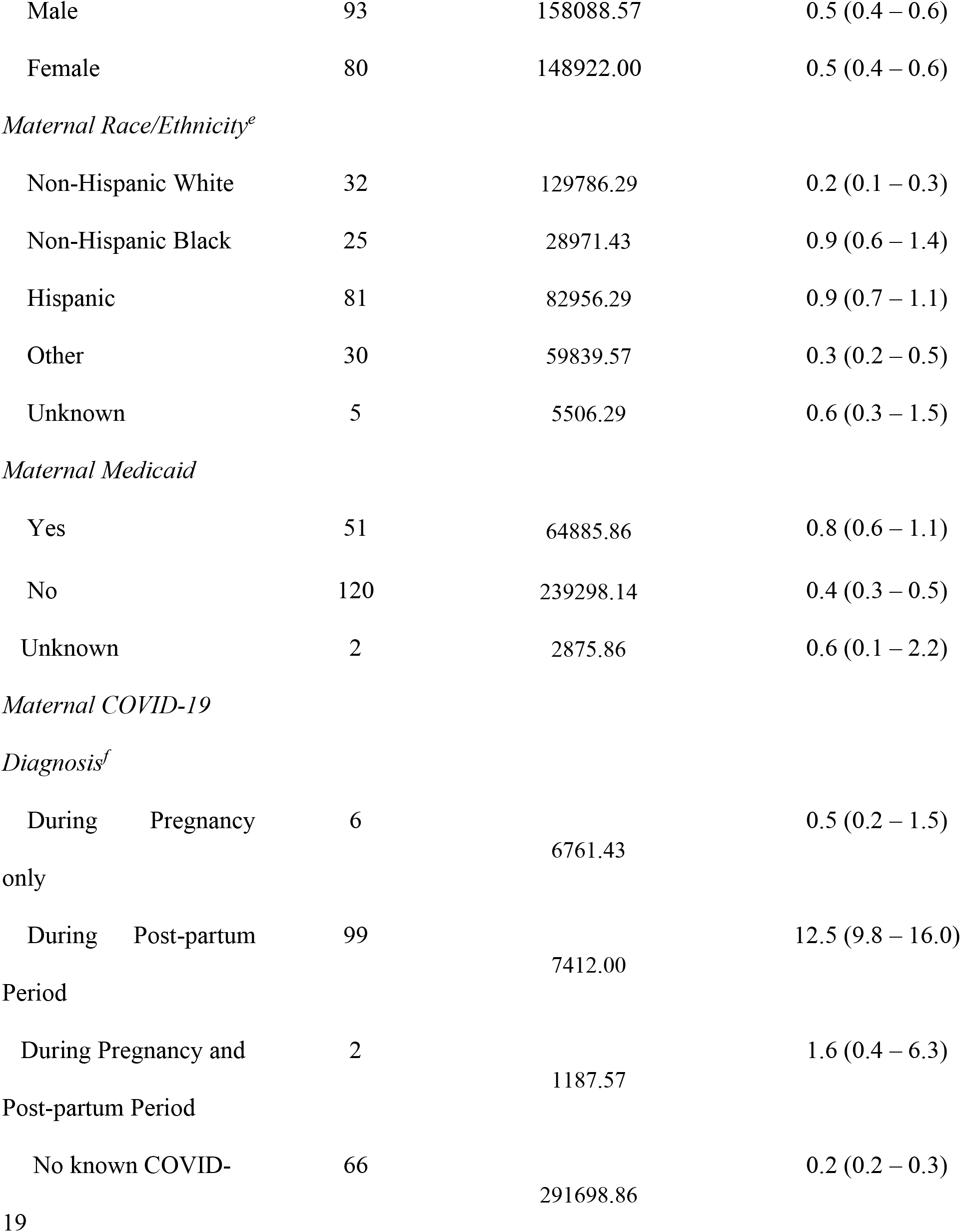
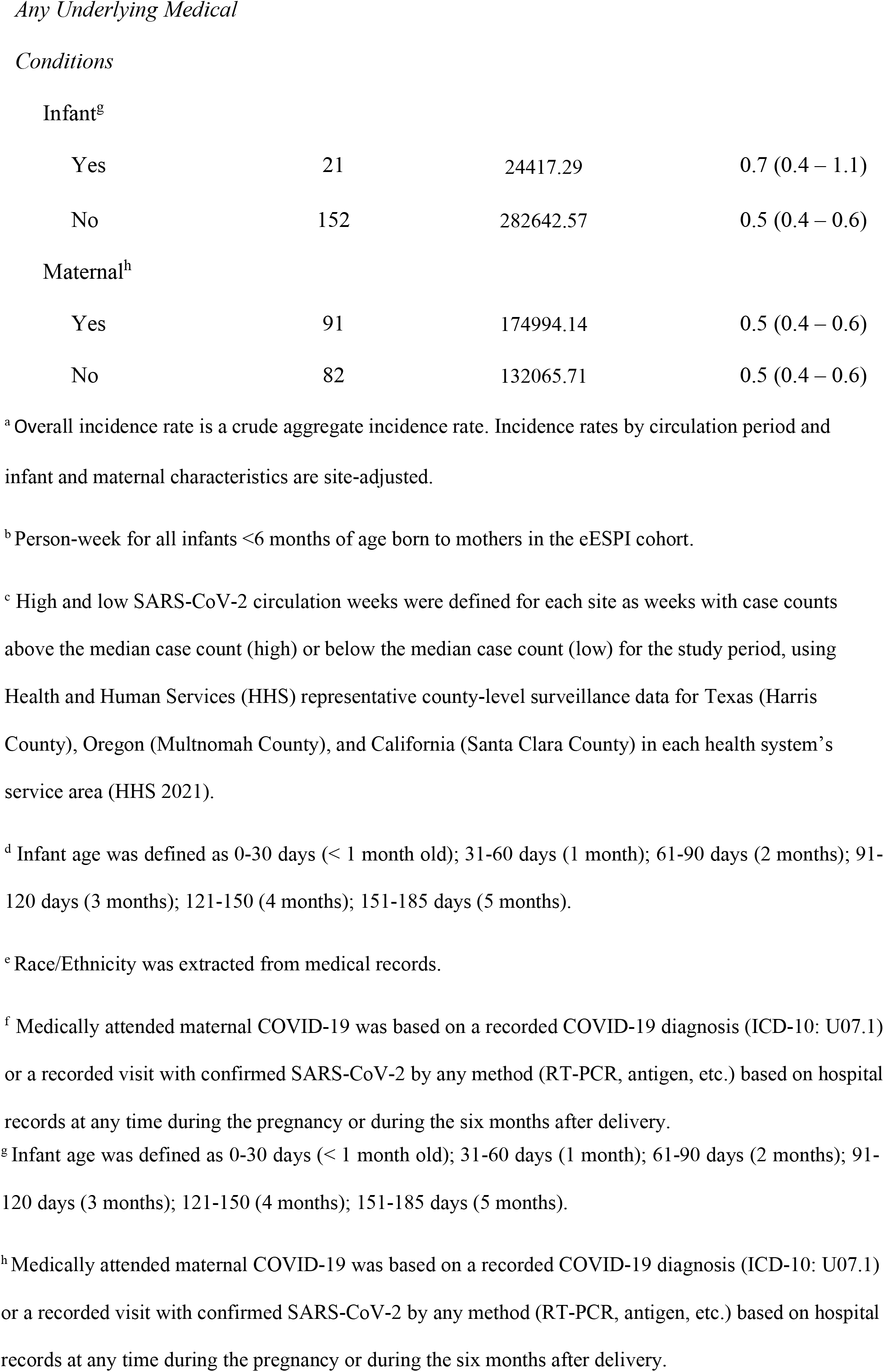
Incidence rates^a^ per 1,000 person-weeks of medically attended COVID-19 episodes among infants under 6 months of age born to mothers in the eESPI cohort, Epidemiology of SARS-CoV-2 in Pregnancy and Infancy (ESPI) Network: Electronic Cohort (eESPI) (N=173).

|  | Infants with<br>Medically<br>Attended<br>COVID-19<br>Episodes | Person-Weeks<sup>b</sup> | Incidence Rates per 1,000<br>Person-Weeks, 95% CI |
| --- | --- | --- | --- |
| Characteristics |  |  |  |
| *Overall incidence rate* | 173 | 307059.86 | 0.6 (0.5 – 0.6) |
| *SARS-CoV-2*<br>*Circulation<sup>c</sup>* |  |  |  |
| Low | 90 | 188198.86 | 0.4 (0.3 – 0.5) |
| High | 83 | 111843.71 | 0.6 (0.5 – 0.8) |
| *Infant Age<sup>d</sup>* |  |  |  |
| Under 1 month | 27 | 67648.00 | 2.0 (1.7 – 2.5) |
| 1 month | 29 | 58250.57 | 2.0 (1.6 – 2.4) |
| 2 months | 28 | 50772.86 | 1.6 (1.3 – 2.1) |
| 3 months | 16 | 45530.00 | 1.3 (0.9 – 1.8) |
| 4 months | 31 | 40628.14 | 1.2 (0.8 – 1.7) |
| 5 months | 42 | 36421.71 | 0.7 (0.4 – 1.2) |
| *Infant Sex* |  |  |  |
| Male | 93 | 158088.57 | 0.5 (0.4 – 0.6) |
| Female | 80 | 148922.00 | 0.5 (0.4 – 0.6) |
| *Maternal Race/Ethnicity<sup>e</sup>* |  |  |  |
| Non-Hispanic White | 32 | 129786.29 | 0.2 (0.1 – 0.3) |
| Non-Hispanic Black | 25 | 28971.43 | 0.9 (0.6 – 1.4) |
| Hispanic | 81 | 82956.29 | 0.9 (0.7 – 1.1) |
| Other | 30 | 59839.57 | 0.3 (0.2 – 0.5) |
| Unknown | 5 | 5506.29 | 0.6 (0.3 – 1.5) |
| *Maternal Medicaid* |  |  |  |
| Yes | 51 | 64885.86 | 0.8 (0.6 – 1.1) |
| No | 120 | 239298.14 | 0.4 (0.3 – 0.5) |
| Unknown | 2 | 2875.86 | 0.6 (0.1 – 2.2) |
| *Maternal COVID-19* |  |  |  |
| *Diagnosis<sup>f</sup>* |  |  |  |
| During Pregnancy only | 6 | 6761.43 | 0.5 (0.2 – 1.5) |
| During Post-partum Period | 99 | 7412.00 | 12.5 (9.8 – 16.0) |
| During Pregnancy and Post-partum Period | 2 | 1187.57 | 1.6 (0.4 – 6.3) |
| No known COVID-19 | 66 | 291698.86 | 0.2 (0.2 – 0.3) |
| Yes | 21 | 24417.29 | 0.7 (0.4 – 1.1) |
| No | 152 | 282642.57 | 0.5 (0.4 – 0.6) |
| Maternal<sup>h</sup> |  |  |  |
| Yes | 91 | 174994.14 | 0.5 (0.4 – 0.6) |
| No | 82 | 132065.71 | 0.5 (0.4 – 0.6) |

<sup>a</sup> Overall incidence rate is a crude aggregate incidence rate. Incidence rates by circulation period and infant and maternal characteristics are site-adjusted.
<sup>b</sup> Person-week for all infants <6 months of age born to mothers in the eESPI cohort.
<sup>c</sup> High and low SARS-CoV-2 circulation weeks were defined for each site as weeks with case counts above the median case count (high) or below the median case count (low) for the study period, using Health and Human Services (HHS) representative county-level surveillance data for Texas (Harris County), Oregon (Multnomah County), and California (Santa Clara County) in each health system's service area (HHS 2021).
<sup>d</sup> Infant age was defined as 0-30 days (< 1 month old); 31-60 days (1 month); 61-90 days (2 months); 91-120 days (3 months); 121-150 (4 months); 151-185 days (5 months).
<sup>e</sup> Race/Ethnicity was extracted from medical records.
<sup>f</sup> Medically attended maternal COVID-19 was based on a recorded COVID-19 diagnosis (ICD-10: U07.1) or a recorded visit with confirmed SARS-CoV-2 by any method (RT-PCR, antigen, etc.) based on hospital records at any time during the pregnancy or during the six months after delivery.
<sup>g</sup> Infant age was defined as 0-30 days (< 1 month old); 31-60 days (1 month); 61-90 days (2 months); 91-120 days (3 months); 121-150 (4 months); 151-185 days (5 months).
<sup>h</sup> Medically attended maternal COVID-19 was based on a recorded COVID-19 diagnosis (ICD-10: U07.1) or a recorded visit with confirmed SARS-CoV-2 by any method (RT-PCR, antigen, etc.) based on hospital records at any time during the pregnancy or during the six months after delivery.

Infants born to mothers with COVID-19 diagnosed during the six-month postpartum period had a higher incidence rate of medically attended COVID-19 episodes (12.5 per 1,000 person-weeks) than infants born to mothers diagnosed with COVID-19 during pregnancy alone (0.5 per 1,000 person-weeks) and compared to infants born to mothers who were never diagnosed with COVID-19 during pregnancy or the postpartum period (0.2 per 1,000 person-weeks).

## DISCUSSION

Among a cohort of >18,000 infants born during the first year of the COVID-19 pandemic in the United States and followed up to six months of age, the overall cumulative incidence of medically attended COVID-19 episodes was 0.95%. Incidence rates varied by level of community transmission, maternal COVID-19 status, maternal race/ethnicity, and infant month of age. The majority of medically attended COVID-19 episodes were outpatient encounters, largely comprising telemedicine visits, and only four infants (<0.1% of the cohort) during their first month of life had COVID-19 associated hospitalizations. Findings from this analysis provide reassurance that the overall risk of medically attended COVID-19 episodes among young infants was low during the study period, which preceded the emergence of both the Delta and Omicron variants, and risk of COVID-19-associated hospitalizations was very low for this group. Incidence rates of medically attended COVID-19 episodes were higher among infants of women with COVID-19 diagnosed during the six-month postpartum period compared to infants born to mothers diagnosed at other times or never diagnosed suggesting potential horizontal transmission between mother and infant and increasing opportunities for community acquisition with increasing infant age. This finding underscores the importance of COVID-19 prevention measures for household members and caregivers of young infants to reduce transmission to young infants. In addition, the highest incidence rates of medically attended COVID-19 episodes were among infants born to Hispanic (0.9 per 1,000 person-weeks) and Non-Hispanic Black mothers (0.9 per 1,000 person-weeks) after adjusting for study site, a finding consistent with those from other studies documenting disparities related to COVID-19 disease burden among minority populations during this study period.^8,9,10,11^ Findings from this analysis suggest that the risk of medically attended COVID-19 episodes among infants in the United States was low during the first year of the pandemic. We were not able to assess the influence of community exposures or preventive practices on incidence rate of medically attended COVID-19 episodes among young infants. However, our finding that incidence rates of medically attended COVID-19 episodes were higher among infants of mothers diagnosed with COVID-19 during the six-month postpartum period suggest that SARS-CoV-2 transmission within households and from caregivers may be important sources of infection for young infants. To reduce the risk of SARS-CoV-2 infection and subsequent medically attended COVID-19 episodes among infants, infection prevention counseling for families and caregivers of young infants should include information about the importance of non-pharmaceutical prevention measures such as handwashing and mask wearing and the benefits of COVID-19 vaccination among eligible persons. Maternal COVID-19 vaccination (either during or prior to pregnancy) may decrease SARS-CoV-2 infection among infants by passive transfer of maternal antibodies to infants and by preventing maternal infections during the postpartum period.^12,13,14,15^ Maternal vaccination during pregnancy has an estimated 61% effectiveness against COVID-19 hospitalization in infants aged <6 months and maternal COVID-19 vaccination is recommended by both CDC and the American College of Obstetricians and Gynecologists (ACOG).^16,17,18^ Other family members and caregivers should also be encouraged to receive COVID-19 vaccine to reduce their risk of SARS-CoV-2 infection and subsequent transmission to young infants in their care.

Infants with medically attended COVID-19 episodes had more underlying medical conditions diagnosed during the first six months of life compared with infants without COVID-19, though this difference was small. This finding is consistent with the literature that found younger children with severe disease were more likely to have underlying conditions.^5,19^ This may be due to increased healthcare seeking behaviors among individuals with medical conditions that afford more opportunities to get SARS-CoV-2 testing.^20^

This analysis has several limitations. First, medically attended COVID-19 episodes were identified based on clinician-initiated testing and/or ICD-10 codes for COVID-19 which may have led to under-ascertainment of episodes and/or differential detection of episodes among infants with different demographic or maternal characteristics. For example, infants of women with COVID-19 diagnosed during the postpartum period may have been more likely to be tested for SARS-CoV-2 because they had a known history of exposure to someone with COVID-19. Second, this analysis was based on data from the first year of the COVID-19 pandemic in the United States and may not be fully generalizable to subsequent periods of SARS-CoV-2 circulation of emerging variants during which younger pediatric age groups were increasingly impacted. Third, this analysis was not able to assess factors that may have led to differences in incidence rates of medically attended COVID-19 episodes by site, such as differences in SARS-CoV-2 circulation patterns, clinical testing practices, and demographic characteristics of the source population. Lastly, the analysis did not account for maternal COVID-19 vaccination status. However, the frequency of women fully vaccinated in this cohort was likely low since women in this cohort were pregnant prior to April 2021 when COVID-19 vaccine became available to the general population of adults <65 years of age in the United States.^21,22^

## CONCLUSIONS

Among a cohort of infants aged <6 months at three sites in the US during the first year of the COVID-19 pandemic, the incidence of medically attended COVID-19 episodes was low with outpatient care encounters accounting for the large majority of episodes. The highest incidence rates of medically attended COVID-19 episodes occurred among infants in the first two months of life and among infants of women diagnosed with COVID-19 during the postpartum period. These findings underscore the importance of COVID-19 prevention measures for pregnant women, household members, and families and caregivers of infants to protect infants who are too young to receive COVID-19 vaccine.

## Data Availability

All data produced in the present study are available upon reasonable request to the authors.

## Data Availability

All data produced in the present study are available upon reasonable request to the authors.

## Abbreviations

COVID-19: coronavirus disease 2019,
SARS-CoV-2: severe acute respiratory syndrome coronavirus 2,
RT-PCR: Reverse Transcription Polymerase Chain Reaction.

## Acknowledgements

We express our gratitude to the Epidemiology of SARS-CoV-2 in Pregnancy and Infancy (ESPI) Network: Electronic Cohort (eESPI) team for their tremendous effort in abstracting electronic health record data. The authors at KPNC thank Andrew K. Hirst for his help with programming and technical support, the authors at KPNW acknowledge Elizabeth Shuster and Jan Van Marter. The authors at Baylor College of Medicine acknowledge Nanette Bond and Patricia Santarcangelo. The authors at Abt Associates acknowledge Michael Duckworth and Peiyi Zhang. The authors from CDC acknowledge Michael Daugherty and Kellye Sliger for data management.

---

**Appendix A:** **Data Dictionary**

| Variable | ESPI Variable | Variable Description | ICD-10 Codes |
| --- | --- | --- | --- |
| Medically Attended Infant COVID-19 | IN_COVID_CONF_VTST# OR IN_ARFI_COVID | Confirmed SARS-CoV-2 by any method for any test for neonate OR Identifies COVID-19 diagnosis code | ICD-10: U07.1 |
| Medically Attended Maternal COVID-19 during Pregnancy | PR_COVID_CONF_VTST# OR PR_ARFI_COVID | Confirmed SARS-CoV-2 by any method for any test during pregnancy OR Identifies COVID-19 diagnosis code during pregnancy | ICD-10: U07.1 |
| Medically Attended Maternal COVID-19 post-partum | PP_COVID_CONF_VTST# OR PP_ARFI_COVID | Confirmed SARS-CoV-2 by any method for any test OR Identifies COVID-19 diagnosis code post-partum | ICD-10: U07.1 |
| Maternal Race/Ethnicity | Calculated based on responses to PR_MAT_ETHNICITY AND PR_MAT_RACE | Maternal race and Maternal ethnicity | No codes |
| Maternal Medicaid | PR_MAT_MEDICAID | Maternal status for Medicaid coverage | No codes |
| Gestational Age | PR_PREG_GA_DEL_NEO# or if blank calculated based on | Gestational age at delivery date | ESPI derived from DELIVERY_DT_NEO#, FIRST_US_DT, GA_FIRST_US |
|  | PR_DELIVERY_DT_NEO | Date of delivery/End of Pregnancy | This should be considered as End of Pregnancy date, ie, if the pregnancy ends in a way other than a delivery (eg, miscarriage, abortion), the date of that outcome should be recorded under this variable. |
|  | PR_EST_CONCEPTION | Estimated date of conception | ESPI calculated from EST_DELIVERY: EST_CONCEPTION = (EST_DELIVERY – 280) |
| Infant Underlying Conditions | IN_HR_CONDITION_PL | High risk medical condition (chronic condition, not pregnancy complication) | See appendix B. |
| Maternal Underlying Conditions | PR_HR_CONDITION_PL | High risk medical condition (chronic condition, not pregnancy complication) | See appendix B. |
| Timing of Maternal COVID-19 | IF PR_COVID_CONF_VTST# OR PR_ARFI_COVID then COVID during pregnancy. |  | ICD-10: U07.1 |
|  | IF PP_COVID_CONF_VTST# OR PP_ARFI_COVID then COVID-19 during post-partum. |  | ICD-10: U07.1 |
| Infant Hospitalization | IN_HOSP_ADM_DT | Hospital admission date |  |
| Infant Death | INF_DEATH | Infant Death |  |
| Live Birth | Calculated based on responses to PR_OUT_STILLBIRTH_NEO# | Stillborn infant (infant born >=20 weeks gestation with no sign of life) [multiple fetuses/neonates] | O60.1; O60.10; O60.12; O60.13; O60.14; P07.21; P07.22; P07.23; P07.24; P07.25; P07.26; P07.3; P07.30; P07.31; P07.32; P07.33; P07.34; P07.35; P07.36; P07.37; P07.38; P07.39 |
|  | PR_PREG_GA_DEL_NEO#/<br>PREG_GA_DEL_EX_NEO/<br>PREG_GA_DEL_ANALYTIC_NEO# | Gestational age at delivery date | Derived or extracted from Medical Records.<br>For Baylor:<br>PREG_GA_DEL_ANALYTIC_NEO# =<br>PREG_GA_DEL_EX_NEO#<br>For KPNW and KPNC:<br>PREG_GA_DEL_ANALYTIC_NEO# =<br>PREG_GA_DEL_NEO# |
|  | PR_OUT_ABORTION_NEO | Induced termination of pregnancy | Z33.2, O04.80. |
|  | PR_OUT_PRETERM_NEO | Preterm birth (A live birth with an estimated gestation <37 weeks) [multiple fetuses/neonates] | O60.1; O60.10; O60.12; O60.13; O60.14; P07.21; P07.22; P07.23; P07.24; P07.25; P07.26; P07.3; P07.30; P07.31; P07.32; P07.33; P07.34; P07.35; |
|  |  |  | P07.36; P07.37; P07.38; P07.39 |
|  | OUT_LIVEBIRTH_NEO | Live birth |  |
|  | PR_OUT_MISCARRIAGE_NEO |  |  |
| Infant Heart<br>*NAC = Non-Acute; AC = Acute | IN_HR_AC_HEART_FAIL_VL | (Acute or chronic) heart failure diagnosis | ICD-10: I50.21, I50.23, I50.31, I50.33, I50.41, I50.43 |
|  | IN_HR_AC_HEART_VL | Acute heart disease diagnosis | ICD-10: I40, J10.82, J11.82, I30, I31.4, R57.0, I50.21, I50.23, I50.31, I50.33, I50.41, I50.43, I50.811, I50.813, I16, I10, I11, I11.9, I11.0, I15, I15.0, I15.9, I16.0, I16.1, I16.9, I20.0, I21, I21.0, I21.1, I21.2, I21.3, I21.4, I21.9, I22, I22.0, I22.1, I22.2, I22.8, I22.9, I24 |
|  | IN_HR_CARD_ARRST_VL | Cardiac arrest diagnosis | ICD-10: I46.9 |
|  | IN_HR_HEART_FAIL_VL | (Acute or chronic) heart failure diagnosis | ICD-10: I50 |
|  | IN_HR_NAC_HEART_VL | Non-acute heart disease diagnosis | ICD-10: A52.0, H35.031-039, I01, I09.2, I05-08, I34, I35, I36, I37, I09, I12, I13, I23, I25-28, I33, I31 (except I31.4), I38, I39, I42-44, I46, I47, I48, I49, I50.1, I50.9, I41, I51, I52, I70, I71, I72, I73, I74, I75, M30.0, M31, I77, I79, I97.0, I97.1, Q21-27, T82, Z95, Q20, Q27.0, Q27.3, Q27.4, Q27.8, Q27.9, Q28, Q89.3, R00.1, Z98.61, Z79.82 |
| Infant Liver<br>*NAC = Non-Acute; AC = Acute | IN_HR_LIVER_VL | (Acute or chronic) Liver disease diagnosis | ICD-10: K70, K73, K74, K75.1-K75.9, I85, K71, K72, K75, K76, K77 |
|  | IN_HR_NAC_LIVER_FAIL_VL | (Non-acute) liver failure diagnosis | ICD-10: K72.1, K76.2 |
|  | IN_HR_AC_LIVER_FAIL_VL | Acute liver failure diagnosis | ICD-10: K72.0, K76.3 |
| Infant Lung | IN_HR_OLUNG_VL | Lung disease diagnosis, not asthma | ICD-10: A15, A17, A18, A19, A31, D86, E84, J40, J41, J42, J43, J47, J60, J61, J62.0, J62.8, J63.6, J64, J67, J68, J69.0, J69.1, J69.8, J70.9, |
|  |  |  | J85, J93, J98, M34, M35, Q34, R09.1, A31.0, B39, B40., B41, B44, B45, B46.0, D86.0, E88.01, J62, J63, J65, J66, J69, J70, J82, J84, J86, J95.0, J96, J98.1, J99, P25, P26, P27, P28, Q33, T86.8, J44 |

**Appendix B:** List of High-Risk Medical Conditions. This set of variables looks at whether each participant had different high-risk medical conditions in diagnosis codes across every visit or admission within the time window for the pull. Each visit or admission for the participant will be evaluated for whether it included one of the listed ICD-10 codes. If any visit/admission for the participant across the entire time window did include one of the ICD-10 codes in question, the variable will be equal to 1 (One or more codes listed). If not, the variable will be equal to 0 (No code listed).

| ESPI Variable | Variable Description | ICD-19 Codes |
| --- | --- | --- |
| IN_HR_EXCLD_ANEMIA_PL <sup>a</sup> | High risk medical condition (chronic condition, not pregnancy complication) for Infant excluding Anemia | Presence of any of the conditions listed below. |
| PR_HR_EXCLD_ANEMIA_PL <sup>b</sup> | High risk medical condition (chronic condition, not pregnancy complication) for Mother during pregnancy excluding Anemia | Presence of any of the conditions listed below. |
| HR_BLOOD_PL | Blood disorder diagnosis | ICD-10: D50.0, D50.8, D50.9, D51, D52, D53, D55-61, D64.0-3, D63.1, D63.0, D63.8, D64.4, D64.8, D64.81, D64.89, D64.9, D66-75, D77 |
| HR_BLOOD_EXCLD_ANEMIA_PL | Blood disorder diagnosis (other than anemia) | ICD-10: D56-61, D64.0-4, D64.8, D63.1, D63.0, D63.8, D64.81, D66-75, D77 |
| HR_DIABT_PL | Diabetes diagnosis | ICD-10: E08-E11 |
| HR_AC_HEART_FAIL_PL | Acute heart failure diagnosis | ICD-10: I50.21, I50.23, I50.31, I50.33, I50.41, I50.43 |
| HR_HEART_FAIL_PL | (Acute or chronic) heart failure diagnosis | ICD-10: I50 |
| HR_CARD_ARRST_PL | Cardiac arrest diagnosis | ICD-10: I46.9 |
| HR_AC_HEART_PL | Acute heart disease diagnosis | ICD-10: I40, J10.82, J11.82, I30, I31.4, R57.0, I50.21, I50.23, I50.31, I50.33, I50.41, I50.43, I50.811, I50.813, I16, I10, I11, I11.9, I11.0, I15, I15.0, I15.9, I16.0, I16.1, I16.9, I20.0, I21, I21.0, I21.1, I21.2, I21.3, I21.4, I21.9, I22, I22.0, I22.1, I22.2, I22.8, I22.9, I24 |
| HR_NAC_HEART_PL | Non-acute heart disease diagnosis | ICD-10: A52.0, H35.031-039, I01, I09.2, I05-08, I34, I35, I36, I37, I09, I12, I13, I23, I25-28, I33, I31 (except I31.4), I38, I39, I42-44, I46, I47, I48, I49, I50.1, I50.9, I41, I51, I52, I70, I71, I72, I73, I74, I75, M30.0, M31, I77, I79, I97.0, I97.1, Q21-27, T82, Z95, Q20, Q27.0, Q27.3, Q27.4, Q27.8, Q27.9, Q28, Q89.3, R00.1, Z98.61, Z79.82 |
| HR_CANCER_PL | Cancer diagnosis | ICD-10: C00, C01, C02, C07, C08, C03, C04, C05, C06, C09, C10, C11, C12, C13, C14, C15, C16, C17, C18, C19, C20, C21, C22, C23, C24, C25, C48, C26, C30, C31, C32, C33, C34, C38, C37, C39, C40, C41, C44, C49, C43, C50, C46, C55, C53, C58, C54, C56, C51, C52, C61, C62, C60, C67, C64, C69, C71, C72, C73, C74, C75, C76, C77, C78, C79, C80, C81, C83, C90, C91, C92, C93, C94, C95, C7A, D02, D37-44, D48, Z51.11, Z51.12, C47, C4A, C57, C65, C66, C68, C70, C7B, D03, Z85, M36.0, Z51.0, Z51.1, C95.00, C95.10, D61.0, D61.2, C82, C84, C85, C86, C88, C96 |
| HR_IMMUNE_PL | Immune disorder diagnosis | ICD-10: B20, B33.3, B59, D73, G70.8, M32.10, M35, M05, M06, R82.8, R84.7, R85.7, R87.7, R89.7, T45, Z21, Z94, B97.3, D47.Z1, D76, M07, M35.0, M35.9, Q89.0, T45.1X1, T86.1, T86.2, T86.3, T86.4, T86.81, T86.85, Z48.2, Z94.0, Z94.1, Z94.2, Z94.3, Z94.4, Z94.82, Z94.83, Z94.84, T86.0, T86.5, D80, D81, D82, D83, D84, D89, D89.81, M08, L40, M32, M30, M33, D46, Z79.5, Z79.52, B97.35, O98.7, D61.9, D73.0 |
| HR_AC_RENAL_FAIL_PL | Acute renal failure diagnosis | ICD-10: N17, N01 |
| HR_NAC_RENAL_FAIL_PL | (Non-acute) Renal failure diagnosis | ICD-10: I13.11, I13.2, K76.7, D63.1, I12.0, I13.11, I13.2, N03, N04, N18, N19, N26.9, N25.0, N25.1, N27.0, N27.1, N27.9, N28.89, Q61, Z49, Z91.15, N02, N06, N07, N08, N11, N14, N15, N16, N25, N26, N28, Q27.1, Q27.2, Q60, Z99.2 |
| HR_AC_LIVER_FAIL_PL | Acute liver failure diagnosis | ICD-10: K72.0, K76.3 |
| HR_NAC_LIVER_FAIL_PL | (Non-acute) liver failure diagnosis | ICD-10: K72.1, K76.2 |
| HR_LIVER_PL | (Acute or chronic) Liver disease diagnosis | ICD-10: K70, K73, K74, K75.1-K75.9, I85, K71, K72, K75, K76, K77 |
| HR_ASTHMA_PL | Asthma diagnosis | ICD-10: J45 |
| HR_OLUNG_PL | Lung disease diagnosis, not asthma | ICD-10: A15, A17, A18, A19, A31, D86, E84, J40, J41, J42, J43, J47, J60, J61, J62.0, J62.8, J63.6, J64, J67, J68, J69.0, J69.1, J69.8, J70.9, J85, J93, J98, M34, M35, Q34, R09.1, A31.0, B39, B40., B41, B44, B45, B46.0, D86.0, E88.01, J62, J63, J65, J66, J69, J70, J82, J84, J86, J95.0, J96, J98.1, J99, P25, P26, P27, P28, Q33, T86.8, J44 |
| HR_OBESE_PL | Obesity diagnosis | ICD-10: E66, O99.210-215, O99.840-845, Z68.4 |
| HR_SEV_OBESE_PL | Severe (morbid) obesity dx | ICD-10: E66.01 |
| HR_OMETBOL_PL | Metabolic disorder diagnosis not obesity | ICD-10: E03-E07, E22, E23, E32, E27, E40-E46, E50-E56, E70-E72, E74, E78, E88, M10, E83, E75.2, E76, E77, E79, E80, E85, E89.1, E89.6, E01, E20, E21, E24, E25, E26, E28, E31, E34 |
| HR_ALT_MENT_PL | Altered mental status diagnosis | ICD-10: F05, F06.0, F06.1, F06.2, F06.30, F06.4, F06.8, G45.4, R40.0, R40.4, R41.82, R44.0, R41 |
| HR_ENCEPHAL_PL | Encephalopathy diagnosis | ICD-10: G93.4, I67.4, K72.90, K72.91 |
| HR_MENINGITIS_PL | Meningitis diagnosis | ICD-10: A02.21, A17.0, A39, A87.0, A87.1, B37.5, B38.4, G00, G01, G02, G03 |
| HR_ANOX_BRN_DMG_PL | Anoxic brain damage diagnosis | ICD-10: G93.1 |
| HR_ENCEPH_MYEL_PL | Encephalitis or myelitis diagnosis | ICD-10: A39.81, G05, G37.3 |
| HR_COMA_PL | Coma diagnosis | ICD-10: R40.20, R40.3 |
| HR_POLYNEURO_PL | Critical illness polyneuropathy diagnosis | ICD-10: G62.81 |
| HR_CEREBRAL_INFARC_PL | Cerebral infarction diagnosis | ICD-10: G43.6, I63.019, I63.119, I63.139, I63.20, I63.219, I63.22, I63.239, I63.30, I63.40, I63.50, I63.59 |
| HR_CEREBRAL_HEMR_PL | Cerebral hemorrhage diagnosis | ICD-10: I60.9, I61.9, I62 |
| HR_CEREBRAL_EDEMA_PL | Cerebral edema diagnosis | ICD-10: G93.6 |
| HR_NEURO_PL | Acute or Chronic Neurologic disease diagnosis | ICD-10: A17, A81, B90.0, F01, F02, F03, F07, F71, F72, F73, E75, G20, G25, G26, G11, M47, G60, G35, G36, G81, G80, G82, G40, G93, G96, I02, I60, I61, I62, I65, I66, I67.82, I67.9, I69, G71.13, R56, T85, I68, A17.0, E75.02, E75.19, E75.4, F84.2, G10, G12-14, G21, G23, G24, G30, G31, G32, G37, G45, G46, G63, G64, G70, G71, G73, G83, G90.3, G91, G94, G95, G99.2, |
|  |  | H49.81, Q05, Q76-79, Q85, Q87.4, Q90-93, Q96, R41, R53.2, R54 |
| HR_AC_NEURO_PL | Only Acute Neurologic disease diagnosis | ICD-10: G04.0, I63, I67.81, I67.82, G45, G45.4, G36, G04, G05, G61.0, I60-I62, G00-G03 |
| HR_ART_VEN_EMBOL_PL | Arterial or venous embolism diagnosis | ICD-10: I74, I76, I82 |
| HR_AC_ENDOCRINE_PL | Acute endocrine complications | ICD-10: E08.1, E08.10, E08.11, E09.1, E09.10, E09.11, E10.1, E10.10, E10.11, E11.1, E11.10, E11.11, E13.1, E13.10, E13.11, E08.0, E08.00, E08.01, E09.0, E09.00, E09.01, E11.0, E11.00, E11.01, E13.0, E13.00, E13.01, E05 |
| HR_AC_GASTRO_PL | Acute gastrointestinal complications | ICD-10: K72.0, B15, B16, B17, B19.0, B19.11, B19.21, K85, J09.X3, J10.2, J11.2 |
| HR_AC_HEMATO_PL | Acute hematological complications | ICD-10: D65, I82.210, I82.290, I82.220, I82.4, I82.6, I82.A1, I82.B1, I82.C1, D76.1, D76.2, D69.3, I26, D57.0, D57.01, D57.02, D57.21, D57.211, D57.212, D57.41, D57.411, D57.412, D57.81, D57.811, D57.812 |
| HR_PULM_EMBOL_PL | Pulmonary embolism diagnosis | ICD-10: I26 |
| HR_CARDITIS_PL | Carditis diagnosis | ICD-10: A39.50, B37.6, I01.0, I01.1, I01.2, I30, I33, I40, I51.4 |
| HR_DIC_PL | Defibrination syndrome diagnosis | ICD-10: D65 |
| HR_KETOACIDOSIS_PL | Ketoacidosis diagnosis | ICD-10: E08.1, E10.1, E13.1 |
| HR_HYPRSMOLRTY_PL | Hyperosmolarity diagnosis | ICD-10: E08.0, E11.0, E13.0 |
| HR_DIABET_COMA_PL | Diabetic coma diagnosis | ICD-10: E08.11, E08.641, E09.11, E09.641, E10.11, E11.01, E11.641, E13.11, E13.641, E15 |
| HR_THYRD_STORM_PL | Thyroid storm diagnosis | ICD-10: E05.01, E05.11, E05.21, E05.31, E05.41, E05.81, E05.91 |
| HR_RHABDOMYOL_PL | Rhabdomyolysis diagnosis | ICD-10: M62.82 |
| HR_MYOSITIS_PL | Myositis diagnosis | ICD-10: M60 |
| HR_MYOPATHY_PL | Critical illness myopathy diagnosis | ICD-10: G72.81 |
| HR_GEST_DIABT_PL | History of gestational diabetes | ICD-10: Z86.32 |
| HR_CHRONHTN_PL | Pregnancy complicated by preexisting hypertension with or without preeclampsia | ICD-10: O10 (O10.0 – O10.99), O11 (O11.1 - O11.9) |
| HR_PREECL_PL | Preeclampsia diagnosis during | ICD-10: O14 (O14.0– O14.05), O14.1 (O14.10– O14.15), O14.2 (O14.20-O14.25), O15 (O15.0- |
|  | pregnancy or postpartum, with or without severe features, and including superimposed preeclampsia | O15.9), O14.9 (O14.90 – O14.95), O11 (O11.1-O11.9) |
| HR_PRCLSEVERE_PL | Preeclampsia with severe features | ICD-10: O14.1 (O14.10– O14.15), O14.2 (O14.20-O14.25), O15 (O15.0-O15.9) |
| HR_HELLP_PL | HELLP syndrome | ICD-10: O14.2 (O14.20-O14.25) |
| HR_GESTHTN_PL | Gestational hypertension (new onset) without proteinuria | ICD-10: O13 (O13.0-O13.9) |

Any underlying medical conditions for the infant included any blood disorder, diabetes, acute or chronic heart failure diagnosis, cardiac arrest, acute or chronic heart disease (congenital), cancer, immune disorder, acute or non-acute renal failure, acute or non-acute liver failure, liver disease, asthma, lung disease (not asthma), altered mental status, encephalopathy, meningitis, anoxic brain damage, encephalitis or myelitis, coma, critical illness polyneuropathy, cerebral infarction, cerebral hemorrhage, cerebral edema, acute or chronic neurologic disease, arterial or venous embolism diagnosis, acute endocrine complications, acute gastrointestinal complications, acute hematological complications, pulmonary embolism, carditis, defibrillation syndrome, ketoacidosis, hyperosmolarity, diabetic coma, thyroid storm, rhabdomyolysis, myositis, or critical illness myopathy. For pregnant women/mothers, additional conditions included history of gestational diabetes, pregnancy complicated by preexisting hypertension with or without preeclampsia, preeclampsia diagnosis during pregnancy or postpartum, with or without severe features, and including superimposed preeclampsia, preeclampsia with severe features, HELLP syndrome, or gestational hypertension (new onset) without proteinuria.

